# Assessing our capability to predict the presence of respiratory diseases at the age of four using data available at one month of age

**DOI:** 10.1101/2021.07.04.21259873

**Authors:** Xinyi Han, Lawrence E. K. Gray, Robert K. Mahar, John B. Carlin, Sarath Ranganathan, Peter J. Vuillermin, Damjan Vukcevic

## Abstract

Chronic respiratory diseases are often difficult to cure and are likely to originate early in life. Therefore, early identification of such diseases is of interest for early prevention.

We explored the potential to predict these almost from birth; using data at 1 month of age, we attempted to predict disease occurrence 4 years later in life. Our data came from the Barwon Infant Study; after cleaning and processing, we had measurements on 41 variables from 401 participants.

We considered three respiratory diseases: asthma, wheeze and hay fever. As predictors, we used a variety of information that would be available in a clinical setting. Of particular interest to our investigation was whether lung function measurements (newly available at such an early age) would helpfully improve predictive accuracy. We also investigated whether maternal smoking (previously associated with respiratory illnesses) is a helpful predictor.

Our methods included logistic regression as the main model, multiple imputation to deal with missing values, stepwise selection and LASSO to select variables, and cross-validation to assess performance. We measured predictive performance using AUC (area under the receiver operating characteristic curve), sensitivity and specificity.

Broadly, we found that the best models had only modest predictive power for each disease. For example, for asthma we achieved an AUC of 0.67, a sensitivity of 68% and a corresponding specificity of 63%. Performance for the other two diseases was similar.

We also found that our lung function measurements did *not* improve predictive performance; somewhat surprisingly, this was also true for maternal smoking. The most useful predictors included, among others, family history of these diseases and variables relating to the size of the infants.

Given the modest performance of these models, our findings suggest that very early prediction of respiratory illnesses is still a challenging task.

## 1 Introduction

### 1.1 Background

Respiratory diseases are one of the challenges facing humanity and cause many deaths and disabilities in the world. Tobacco smoke (Vanker et al., 2017), indoor air pollution from burning fuels, air pollution from transportation and industrial sources (Brauer, 2010; Ferkol & Schraufnagel, 2014) are considered to be the cause of most respiratory diseases. As air pollution gets worse, the number of people suffering from chronic respiratory diseases increases every year. Even though a lot of effort is spent on chronic respiratory diseases every year, there is no complete cure for chronic respiratory diseases. The report from the Forum of International Respiratory Societies demonstrates the link between respiratory diseases and the environment and highlights the importance of preventing respiratory diseases from before birth (Forum of International Respiratory Societies, 2017).

Most chronic respiratory diseases start developing early in life (Carraro et al., 2014; Stocks et al., 2013), are incurable, and can last throughout a person’s life. Chronic respiratory diseases are a variety of chronic infections of the lungs and airways that have many adverse effects on people’s daily lives. Childhood is an important time for healthy development, and many chronic respiratory diseases begin in infancy or in utero (Manuck et al., 2016). Because the lungs are not fully developed during childhood, they are highly susceptible to environmental influences (Carraro et al., 2014). Poor lung development can lead to lifelong defects in the respiratory system that affect people throughout their lives.

The diagnosis and treatment of respiratory diseases are complex, especially in developing countries. Early detection of disease and prediction of the likelihood of children developing respiratory disease can reduce mortality. While chronic disease cannot be cured, it is treatable and morbidity can be prevented (Ferkol & Schraufnagel, 2014). From a prevention perspective, predicting those at increased risk of developing chronic respiratory disease allows for interventions that not only prevent the progression of the disease, but also help patients maintain lung function and quality of life. Therefore, if allergies and asthma in children can be detected early and treated effectively, we may be able to prevent them from becoming chronic and serious diseases in adulthood. A model for predicting chronic respiratory disease in children is helpful for this purpose.

### 1.2 Development of, and risk factors for, respiratory diseases

In an effort to prevent and treat respiratory diseases as early as possible, a large and growing body of literature investigates the origins of disease development (Elo & Preston, 1992). It is undisputed that early abnormal development of organ systems has long-term effects on an individual’s health. It is important to know when respiratory diseases begin to develop, and understanding the origins of respiratory diseases is useful in order to select the features that would help to predict such diseases.

As stated in the Introduction, respiratory disease usually begins early in life. A respiratory disease may be present for several years before the person is diagnosed with it. This means that respiratory disease develops in a state that is unrecognised by the individual. Asthma, the most prevalent chronic disease of childhood, is very commonly under-diagnosed in Australia. In studies with 2,523 South Australian adults as a random sample, with a consistent definition of asthma, 19.2% of those with asthma were undiagnosed (Aaron et al., 2018).

Smoke exposure in utero is one of the best known risk factors for decreased lung function and respiratory disease in infants. Maternal smoking during pregnancy has a high potential to negatively affect the development of the infant’s lungs and increase the risk of asthma and wheezing during childhood (Gilliland et al., 2001; Stick et al., 1996; Carlsen et al., 1997; Hanrahan et al., 2012; Weitzman et al., 1990; Martinez et al., 1995). A study with a sample of 500 newborn Australian infants illustrates the relationship between smoke exposure in utero, family history of asthma, maternal hypertension, and low lung function in newborns (Stick et al., 1996).

There is evidence of an association between poor lung function in newborns and the presence of later respiratory diseases. In a prospective survey of 1,246 children followed at three and six years of age, it was shown that poorer lung function at birth can lead to transient wheeze, but symptoms of wheeze are not associated with an increased risk of asthma in six-year-olds or later in life (Martinez et al., 1995). However, the same study also shown there is a relationship between the early onset of symptoms of wheeze and the development of asthma. Another study came to a similar conclusion that the presence of wheeze in one- and three-year-old children is associated with low lung function early in life (Pike et al., 2011).

In addition to these factors, as was found in other studies, family history of respiratory diseases, IgE, birth weight, siblings and daily care are all risk factors for respiratory diseases (Steffensen et al., 2000; Stick et al., 1996; Eysink et al., 2005; Brims & Chauhan, 2005).

### 1.3 The current state of respiratory disease prediction

If children with high levels of asthma can be identified early in life using readily available clinical parameters, then timely prevention and treatment of these children can be implemented. Numerous studies (Schenker et al., 1983; Eysink et al., 2005; Gergen, 2001) are exploring the risk factors that contribute to respiratory diseases, and there are many studies (Caudri et al., 2009; Grabenhenrich et al., 2015; Keller et al., 2017) using different predictors to obtain predictive models to predict respiratory diseases.

A study used data from 654 children aged 1 to 4 years who had coughing for more than 5 consecutive days to develop an asthma prediction model that attempted to predict the probability of having asthma at age 6. The model used age, wheezing, family history of pollen allergy, and IgE test results as predictors, and the accuracy of the prediction model was improved with the addition of the IgE test (Eysink et al., 2005). However, this model is difficult to apply to the primary care situation because of the need for allergen testing. The predictive power of the model for other populations is unclear because the data used to validate the model consisted of children who sought medical attention because of asthma symptoms.

Similarly, Caudri et al. (2009) developed a child-specific prediction model using 2171 children aged 0 to 4 years with asthma-like symptoms to predict the probability of having asthma at age 7–8 years. The model had good predictive power for children with wheezing or coughing symptoms, but was not suitable for all children, as the predictive power for asymptomatic children is not known.

Much less research has been done on the predictors and risks of hay fever than on asthma and wheezing, but it is also a very prevalent chronic disease (Grabenhenrich et al., 2015; Omenaas et al., 2008). Grabenhenrich et al. (2015) explored the relationship between early environment and lifestyle and hay fever using data on the environmental, behavior and allergy histories of 1,314 children from birth to 20 years of age. The study shown that although there is evidence that early living environment and lifestyle are associated with an increased risk of hay fever, they do not predict the occurrence of hay fever. However, early food allergies and parental hay fever could serve as predictors.

Available evidence suggests that factors such as early life environment (Colley & Holland, 1967), especially maternal smoking (Carlsen et al., 1997; Hanrahan et al., 2012; Weitzman et al., 1990; Martinez et al., 1995), lung function, family history of respiratory diseases (Stick et al., 1996), and the onset of wheezing symptoms are all associated with later respiratory diseases. However, few predictive models are clinically applicable, and some predictive models use selected samples that cannot be applied to all children. Thus more exploration of respiratory diseases is needed.

### 1.4 Measuring lung function

The multiple breath washout (MBW) technique has been used many times to understand the development of respiratory disease in childhood. It was originally designed to measure uneven gas distribution in the lungs, but as devices have advanced, the technique has been improved to measure functional residual capacity (FRC), lung clearance index (LCI) and cumulative expired volume (CEV) during tidal breathing (Fowler et al., 1952). This technique has been adapted for use in unsedated newborns, which will help to understand the lung function of the newborn (Sinhal et al., 2010). These three measurements are important indicators as one of the purposes explored in this study.

### 1.5 Data

The data used in this paper are from the Barwon Infant Study (BIS), which aims to advance our understanding of the mechanisms underlying non-communicable diseases. In the Barwon region of southeastern Australia, which has similar demographic characteristics to the rest of Australia, women were recruited at their first antenatal hospital visit. A total of 1,064 women and 1,074 infants were enrolled into the study. Subsequent reviews with mothers and infants were conducted at fixed intervals before and after birth, with the infant’s reviews at age four ending at the end of 2014. Parental and infant data were collected at different times by means of questionnaires and clinical examination, and infant lung function data were collected at the age of one month (Vuillermin et al., 2015).

### 1.6 Research goals

The aim of this study was to assess the ability to predict respiratory diseases in 4-year-old children using information obtained 1 month after birth and earlier in the BIS population. Specifically, the purpose of this study is to develop a predictive model by selecting logistic regression models to investigate the information that is important for accurate prediction in a clinical-based setting. And based on previous literature, we will explore the ability of three indicators of MBW (LCI, FRC and CEV) and maternal smoking in predicting respiratory phenotypes.

## 2 Data and methods

### 2.1 Variables

As described in Section 1.5, the dataset for this study comprised data collected by BIS from Barwon district and its surrounding areas. To begin with, the variables of interest were selected from the BIS dataset based on previous studies and available data. The obtained dataset contained 1074 participants and 2685 variables, excluding the three lung function measurement variables (LCI, FRC and CEV). However, we found a large number of missing values in this dataset and further selection of variables was required. The process of further selection of participants and variables will be explained below.

LCI, FRC, and CEV were necessary for one of the purposes of this study because they are data that measure lung function in 1-month-old infants. First we needed to convert the MBW data, there were a total of 1,197 MBW records and a total of 432 infants (including 4 pairs of twins) had their lung function measurement data completely collected and recorded. This relates to the fact that one infant can have multiple lung function measurements. This needs to be addressed before the data can be properly analyzed, by summarising the information from multiple measurements into a single set of measurements. Of the infants for whom MBW records exist, there are between 1 and 7 recordings per individual for each lung function parameter. The same measurement in each participant was averaged so that each child have only one index and one row containing three variables (LCI,FEC and CEV). And merge this data set with the above data set that has 1,074 rows, keeping the complete row of MBW data.

After excluding participants with missing values for the lung function variables, 31 of the remaining participants had missing values for all outcomes and were also excluded. Of the 2,688 variables, the total proportion of missing values was 72.5% due to the large number of missing values for most variables. Of these, Figure 1 shows that 1,986 variables have more than 50% of their values missing which are distrusted in the dataset (Steyerberg et al., 2019); all of these variables were excluded from further analyses.

**Figure 1:**
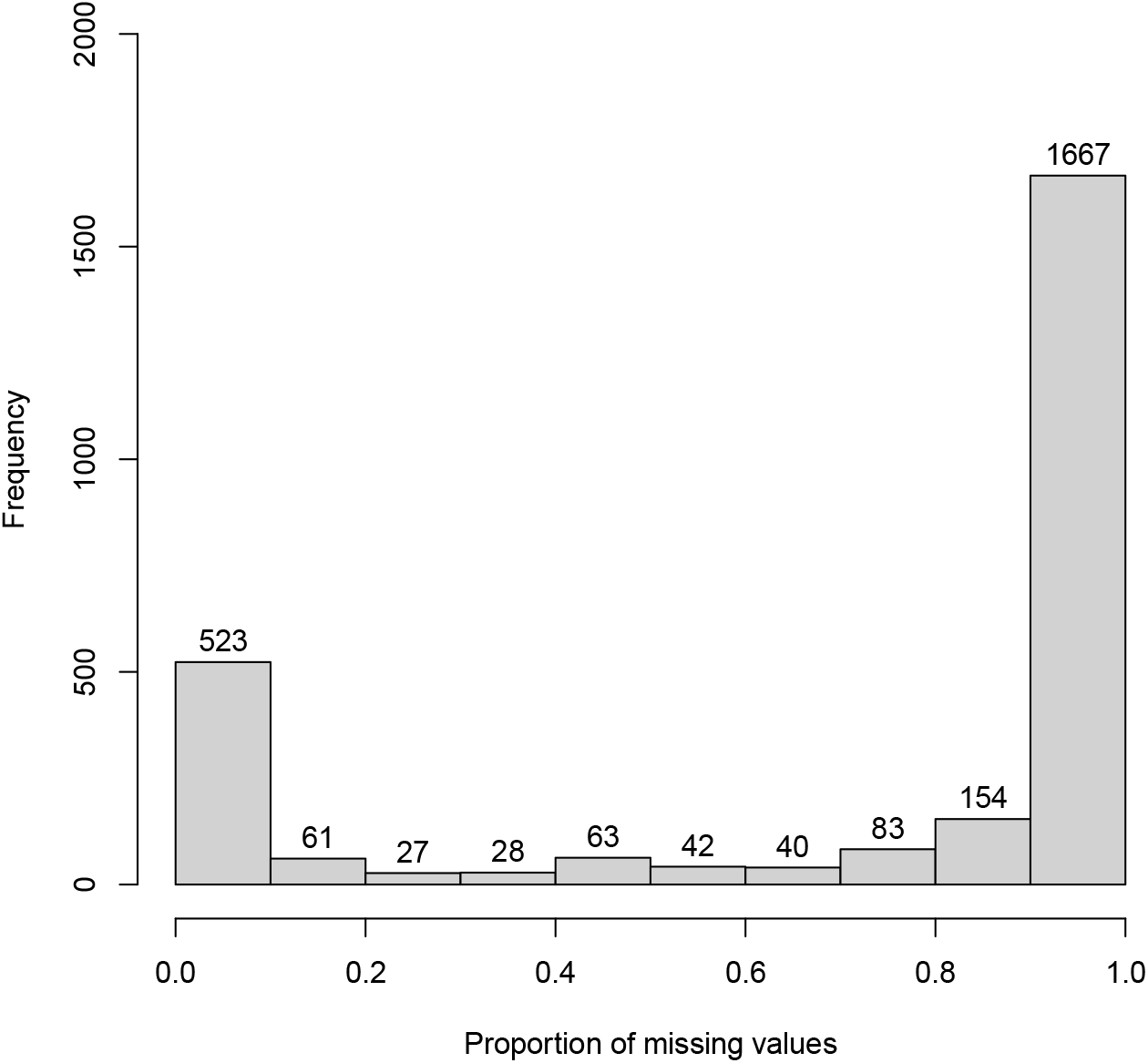
Distribution of the proportion of missing values of variables.

Table 2 lists the final selected variables for analysis and their descriptions. These include infant lung function data, anthropometric data at one month of age, demographics, the child’s environmental exposures, and parental and infant diets for a total of 38 predictor variables. The baseline characteristics of these variables are presented in the Table 3.

**Table 1:**
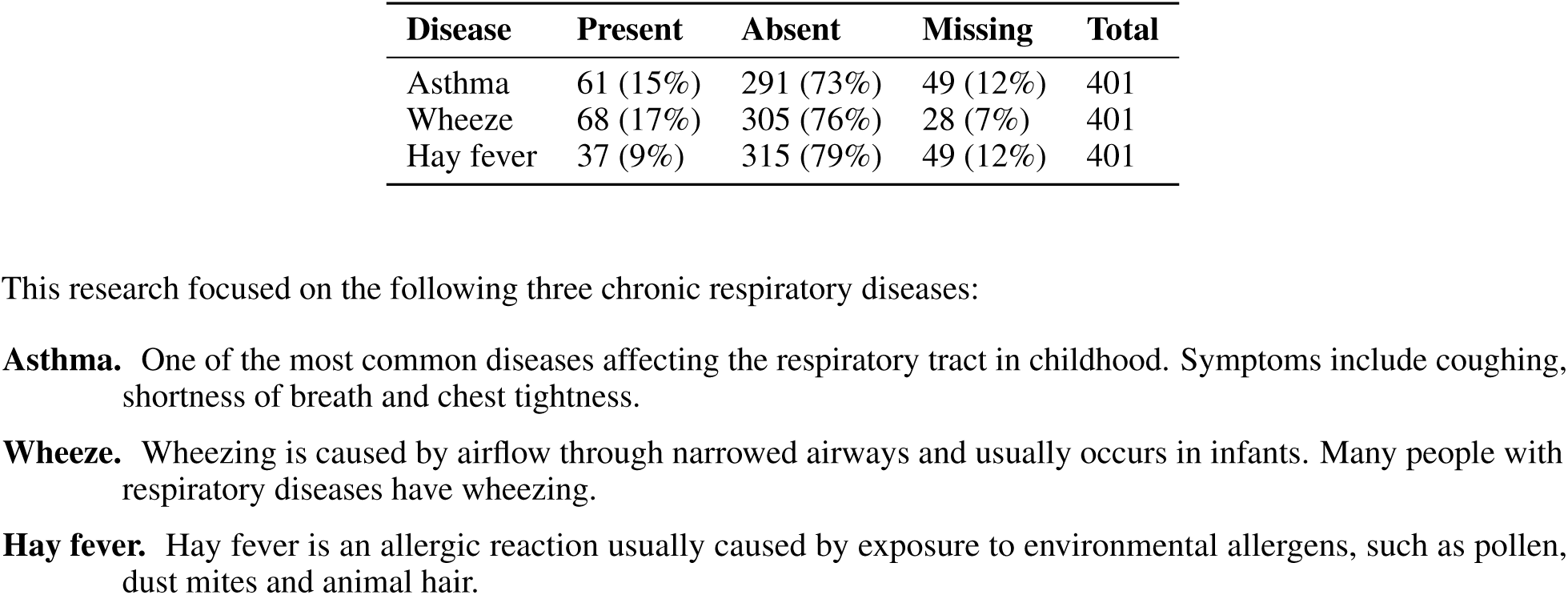
Summary of outcomes.

| Disease | Present | Absent | Missing | Total |
| --- | --- | --- | --- | --- |
| Asthma | 61 (15%) | 291 (73%) | 49 (12%) | 401 |
| Wheeze | 68 (17%) | 305 (76%) | 28 (7%) | 401 |
| Hay fever | 37 (9%) | 315 (79%) | 49 (12%) | 401 |

This research focused on the following three chronic respiratory diseases:
**Asthma.** One of the most common diseases affecting the respiratory tract in childhood. Symptoms include coughing, shortness of breath and chest tightness.
**Wheeze.** Wheezing is caused by airflow through narrowed airways and usually occurs in infants. Many people with respiratory diseases have wheezing.
**Hay fever.** Hay fever is an allergic reaction usually caused by exposure to environmental allergens, such as pollen, dust mites and animal hair.

**Table 2:** Definitions of the variables.

| Variable | Description |
| --- | --- |
| *Variables for 1 month after birth or earlier* |  |
| LCI | Lung clearance index |
| FRC | Functional residual capacity |
| CEV | Cumulative expired gas volume |
| Thc_b | Thigh circumference at birth |
| Abc_b | Abdominal circumference at birth |
| Uparm_b | Middle upper arm circumference at birth |
| Hc_b | Head circumference at birth |
| Wt_b | Weight at birth |
| Trc_b | Tricep skin fold at birth |
| Subfold_b | Subscapular fold at birth |
| Len_b | Length at birth |
| Mage | Mother's age at conception |

Table 2: Definitions of the variables.
| Variable | Description |
| --- | --- |
| Fage | Father's age at conception |
| Meducation | Mother's highest education (1 = Less than year 10; 2 = Year 10 or equivalent; 3 = Year 12 or equivalent; 4 = Trade/Certificate/Diploma; 5 = Bachelor degree; 6 = Postgraduate degree; 7 = Other) |
| Feducation | Father's highest education (1 = Less than year 10; 2 = Year 10 or equivalent; 3 = Year 12 or equivalent; 4 = Trade/Cert/Dip; 5 = Bachelor degree; 6 = Postgraduate degree; 7 = Other) |
| Siblings | Any siblings (0= No; 1 = Yes) |
| Hhincome | Household income last 12 months (1 = Less than 25,000; 2 = 25,000 to 49,999; 3 = 50,000 to 74,999; 4 = 75,000 to 99,999; 5 = 100,000 to 149,000; 6 = More than 150,000) |
| Pets | Pet ownership (0 = No; 1 = Yes) |
| Livestocks | Livestock ownership (0 = No; 1 = Yes) |
| Fh_asthma | Family history of asthma (0 = No; 1 = Yes) |
| Fh_eczema | Family history of eczema (0 = No; 1 = Yes) |
| Fh_hayfever | Family history of hayfever (0 = No; 1 = Yes) |
| Mpsmoke | Mother smoking during pregnancy (0 = Non pre,T1,T2,T3; 1 = Light pre, none T1,T2,T3; 2 = Mod pre, none T1,T2,T3; 3 = Light pre or T1, none T2,T3; 4 = Mod pre or T1, none T2,T3; 5 = Smoker pre,T1,T2,T3) <sup>a</sup> |
| Anympsmoke | Any smoking by the mother during pregnancy (0 = No; 1 = Yes) |
| Passive_p12 | Any passive smoking pre,T1,T2 (0 = No; 1 = Yes) |
| Milk | Milk (ml)/day (0 = none 1 = <250 ml/day 2 = 250 to 500 ml/day 3 = 500 to 750 ml/day 4 = 750+ ml/day) |
| Egg | Eggs/week (0 = don't eat eggs; 1 = <1 egg/week; 2 = 1–2 eggs/week; 3 = 3–5 eggs/week; 4 = 6+ eggs/week) |
| Teaone | Never drank tea (0 = not ticked; 1 = ticked) |
| Cofnone | Never drank coffee (0 = not ticked; 1 = ticked) |
| Hygwashhn | Baby handwash freq (0 = Not at all; 1 = 1–2 times; 2 = 3–4 times; 3 = 5+ times) |
| Hygbathsh | Baby wash freq (1 = Hardly ever; 2 = Once a wk; 3 = Several times a wk; 4 = Once every day; 5 = More than once a day) |
| Slppos | Sleep position (1 = Side; 2 = Stomach, face down; 3 = Stomach, face side; 4 = Back, face up; 5 = Back, face side; 6 = No usual position) |
| Cc | NonParent childcare (0 = No; 1 = Yes) |
| Osidehrs | Outside Hrs/W |
| Vacc | Baby vaccines (0 = No; 1 = Yes) |
| Pest | Pesticides (0 = No; 1 = Yes) |
| Chlhm | Chlorine freq (1 = Every day; 2 = A few times a week; 3 = About once a week; 4 = Less than once a week; 5 = 1–3 times a month; 6 = Not at all) |
| Chlbabyb | Chlorine in babybed (0 = No; 1 = Yes) |
| *Variables for 4 years of age* |  |
| asthma_4y | Asthma at 4 years of age (0 = No; 1 = Yes) |
| wheeze_4y | Wheeze at 4 years of age (0 = No; 1 = Yes) |
| hayfever_4y | Hay fever at 4 years of age (0 = No; 1 = Yes) |

<sup>a</sup> Pre, T1, T2 and T3 represent different periods of pregnancy. Pre: preconception; T1: First trimester (conception to 12 weeks); T2: Second trimester (12 to 24 weeks); T3: Third trimester (24 to 40 weeks).

**Table 3:** Summary of variables. For continuous variables, we show the mean and standard deviation (the latter is shown in parentheses). For categorical variables, counts and percentages (shown in parentheses) are shown.

| Variable | Summary |
| --- | --- |
| LCI | 6.087 (0.39) |
| FRC | 127.5 (21.75) |
| CEV | 771.6 (114.89) |
| Thc_b | 13.52 (1.71) |
| Abc_b | 30.28 (3.04) |
| Uparm_b | 9.819 (1.21) |
| Hc_b | 34.75 (1.56) |
| Wt_b | 3.527 (0.51) |

| Variable | Summary |
| --- | --- |
| Trc_b | 4.936 (1.22) |
| Subfold_b | 4.796 (1.18) |
| Len_b | 50.93 (2.29) |
| Mage | 31.57 (4.71) |
| Fage | 33.34 (5.45) |
| Meducation |  |
| Less than year 10 | 3 (0.75%) |
| Year 10 or equivalent | 16 (4%) |
| Year 12 or equivalent | 64 (16%) |
| Trade/Cert/Dip | 102 (25.5%) |
| Bachelor degree | 146 (36.5%) |
| Postgraduate degree | 69 (17.25%) |
| Feducation |  |
| Less than year 10 | 8 (2.05%) |
| Year 10 or equivalent | 21 (5.38%) |
| Year 12 or equivalent | 70 (17.95%) |
| Trade/Cert/Dip | 156 (40%) |
| Bachelor degree | 96 (24.62%) |
| Postgraduate degree | 39 (10%) |
| Siblings | 238 (59.35%) |
| Hhincome |  |
| Less than 25,000 | 7 (1.79%) |
| 25,000 to 49,999 | 37 (9.44%) |
| 50,000 to 74,999 | 73 (18.62%) |
| 75,000 to 99,999 | 90 (22.96%) |
| 100,000 to 149,000 | 142 (36.22%) |
| More than 150,000 | 43 (10.97%) |
| Pets | 305 (76.44%) |
| Livestocks | 29 (7.32%) |
| Fh_asthma | 212 (54.08%) |
| Fh_eczema | 186 (47.57%) |
| Fh_hayfever | 251 (64.52%) |
| Mpsmoke |  |
| Non pT,T1,T2,T | 340 (86.29%) |
| Light pT, none T1,T2,T3 | 18 (4.57%) |
| Mod pre, none T1,T2,T3 | 3 (0.76%) |
| Light pTorT1, none T2,T3 | 9 (2.28%) |
| Mod pTorT1, none T2,T3 | 3 (0.76%) |
| Smoker pT,T1,T2,T3) | 21 (5.33%) |
| Passive_p12 | 44 (11.28%) |
| Anympsmoke | 54 (13.71%) |
| Milk |  |
| none | 10 (2.58%) |
| <250 ml/day | 112 (28.94%) |
| 250 to 500 ml/day | 196 (50.65%) |
| 500 to 750 ml/day | 57 (14.73%) |
| 750+ ml/day | 12 (3.10%) |
| Egg |  |
| Don't eat eggs | 16 (4.14%) |
| <1 egg/week | 93 (24.09%) |
| 1–2 eggs/week | 164 (42.49%) |
| 3–5 eggs/week | 101 (26.17%) |
| 6+ eggs/week | 12 (3.11%) |
| Teanone | 155(40.16%) |
| Cofnone | 208(53.89%) |
| Hygwashhn |  |
| Not at all | 159 (40.05%) |
| 1–2 times | 211 (53.15%) |
| 3–4 times | 16 (4.03%) |

| Variable | Summary |
| --- | --- |
| 5+ times | 11 (2.77%) |
| Hygbathhn |  |
| Hardly ever | 3 (0.75%) |
| Once a wk | 15 (3.76%) |
| Several times a wk | 295 (73.93) |
| Once every day | 85 (21.30%) |
| More than once a day | 1 (0.25%) |
| Slppos |  |
| Side | 13 (3.27%) |
| Stomach, Face Side | 8 (2.01%) |
| Back, Face Up | 46 (11.56%) |
| Back, Face Side | 326 (81.91%) |
| No Usual Position) | 5 (1.26%) |
| Cc | 19 (4.76%) |
| Osidehrs | 26.59 (121.38) |
| Vacc | 336 (84.63%) |
| Pest | 152 (38.09%) |
| Chlhm |  |
| Every day | 5 (1.26%) |
| A few times a week | 9 (2.27%) |
| About once a week | 66 (16.62%) |
| Less than once a week | 29 (7.31%) |
| 1–3 times a month | 60 (15.11%) |
| Not at all | 228 (57.43%) |
| Chlbabyb | 6 (1.54%) |

There were 401 study participants in the above selected dataset. The three outcomes (`asthma_4y`, `wheeze_4y` and `hayfever_4y`) indicate the respiratory health status of the participants at the age of four years, i.e. whether or not the observer had any of the three diseases. Of these, 61 had asthma at four years of age, 68 had wheeze in the past 12 months at four years of age, and 37 had hay fever. Table 1 provides the sample size and distribution for each outcome.

### 2.2 Missing data

The dataset is composed of 41 variables and 401 participants. The proportion of missing values for each column of variables is summarized in Figure 2 The proportion of missing values in each column does not exceed 4%, which is acceptable. The visualisation of the missing value pattern is provided. These missing values were imputed using multiple imputation (MI).

**Figure 2:**
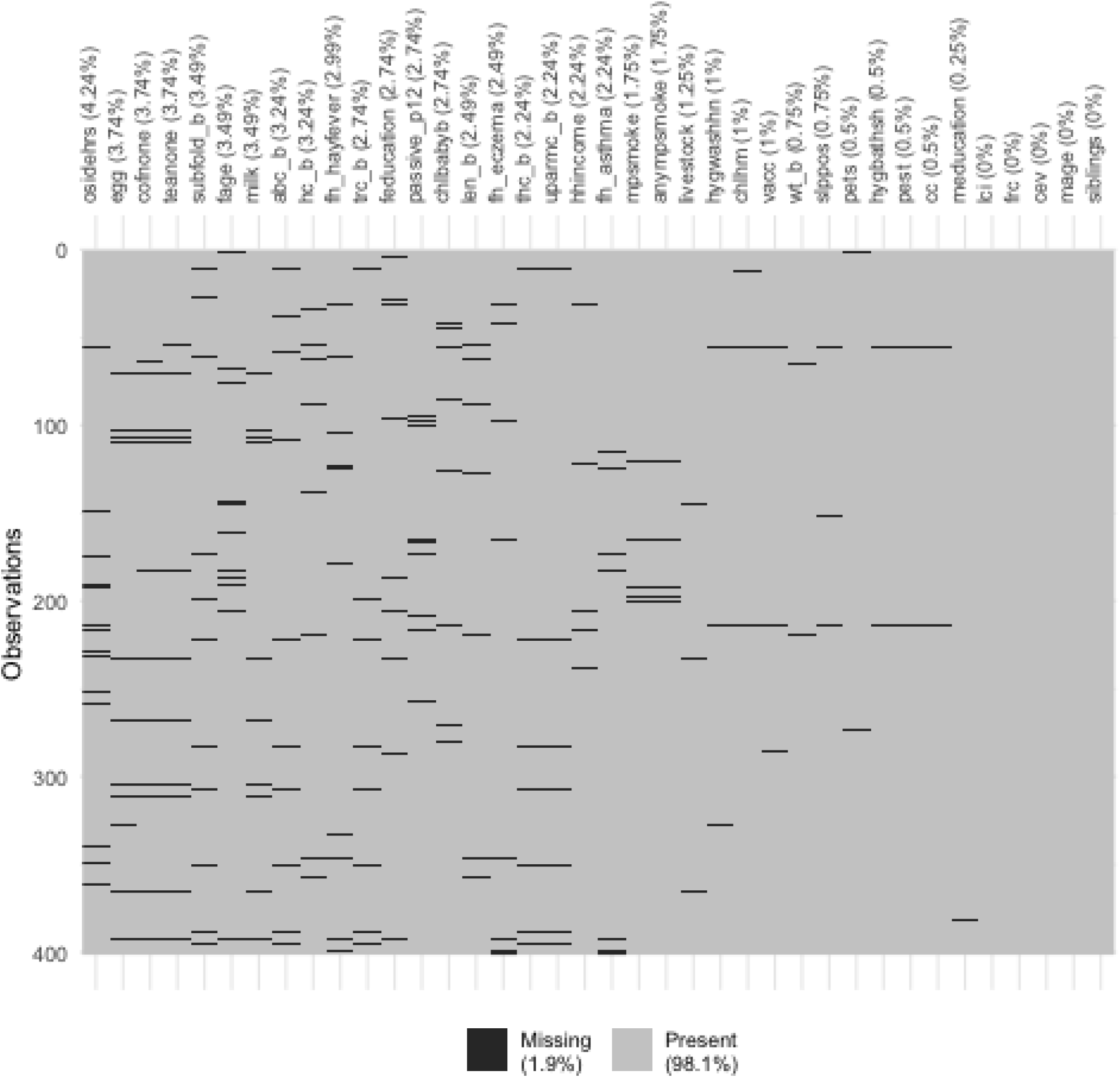
Visualisation of the missing values.

We used the MI approach known as Multivariate Imputation by Chained Equations (MICE), as implemented by the `mice` R package (Buuren & Groothuis-Oudshoorn, 2010).

We imputed the dataset with 10 iterations of Gibbs sampling and set the imputation number to 5 (*m* = 5), while the built-in imputation model used was Predictive Mean Matching (PMM) to estimate missing values. The PMM is a semi-parametric imputation method (Buuren & Groothuis-Oudshoorn, 2010) where the value closest to the predicted value of the missing value is selected from a range of observations to fill in the missing value. In using this MI algorithm, we have assumed that the missing values are missing at random (MAR), a commonly used assumption and one that we think is reasonable here.

After imputation, we had 5 complete datasets consisting of observations and imputed values, including 38 predictors and 401 observations for 3 outcomes.

Note that, in the data analysis for any given disease, we excluded participants with missing values for this disease (i.e. missing values in the outcome variable). An alternative approach is to use the imputed values for this disease; we did not attempt this.

### 2.3 Data analysis

For all of our analyses, we used a logistic regression model with various choices of predictor variables. Unless otherwise stated, we used all available variables as predictors. We performed the same analyses for each of the three diseases, with each one in turn used as the response variable.

We used two different methods for selecting variables: stepwise selection and LASSO. We applied both for each model and compared results.

The `stepAIC()` function from MASS was used to implement stepwise selection, applying forward selection and backward elimination, and the function eventually returns the best model for each dataset.

Fitting a logistic regression model using multiple imputed datasets is slightly different from fitting a single dataset. For each outcome, after applying stepwise selection to each of the five complete datasets to select models, five less identical models were obtained. To fit the logistic regression model using the same model, the five results were analysed. A list of variables that appeared at least half the time (i.e. three times) was selected and these variables were then used to fit logistic regression model in each dataset.

For LASSO, we use the `train()` function from the `caret` R package, where we apply CV to the function to find the best tuning parameter to minimize test error and get the results of fitting the model to all the dataset using this parameter for each dataset. We end up with a list of five variables selected by the LASSO. The final model is selected in the same way as stepwise selection, by identifying variables that show up more than three times.

We attempted to explore how useful the lung function and smoking variables were by looking at the models with only these variables and what had happened when these variables were added to the best set of variables obtained earlier.

### 2.4 Software

All data preprocessing, including creating subsets, feature selection, and handling of missing values, was done in RStudio (Version 1.3.1073) using the R programming language (R Core Team, 2013). The following R packages were used: `dplyr` for manipulating the dataset, `naniar` for missing value visualisation, `mice` for imputing missing values, caret for choosing models and fitting logistic regression model, and `MLeval` for ploting ROC curves.

## 3 Results

We applied our modelling approach to each of the three diseases, asthma, hay fever and wheeze. We present our results for each disease in turn. For clarity of presentation, we have given specific names to each of the ‘best’ models for each disease, as shown in Table 4).

**Table 4:** Definition and naming of models.

| Outcome | Model name | Description |
| --- | --- | --- |
| Asthma | Model 1 | Model selected by stepwise selection when the response variable is asthma. |
|  | Model 2 | Model selected by LASSO when the response variable is asthma. |
| Wheeze | Model 3 | Model selected by stepwise selection when the response variable is wheeze. |
|  | Model 4 | Model selected by LASSO when the response variable is wheeze. |
| Hay fever | Model 5 | Model selected by stepwise selection when the response variable is hay fever. |
|  | Model 6 | Model selected by LASSO when the response variable is hay fever. |

### 3.1 Asthma

#### 3.1.1 Using variable selection

Table 5 shows the list of variables arising from stepwise selection, while Table 6 shows those selected by LASSO. As we can see, there are some differences in the variables selected by the two methods. Each table also shows the average coefficients of the variables in each model (averaged across imputed datasets). For variables that are present in both models, the coefficients in the two models are very close to each other. In particular, children with a family history of asthma and hay fever have an increased risk of asthma, and increasing time spent outside reduces the risk of the disease.

**Table 5:** Estimated coefficients for Model 1.

| Variable | Coefficient |
| --- | --- |
| Tricep skin fold at birth | 0.28 |
| Abdominal circumference at birth | −0.07 |
| Family history of asthma | 0.56 |
| Family history of hay fever | 0.78 |
| Baby vaccines | 0.97 |
| Outside hours | −0.07 |

**Table 6:** Estimated coefficients for Model 2.

| Variable | Coefficient |
| --- | --- |
| LCI | −0.36 |
| Tricep skin fold at birth | 0.26 |
| Abdominal circumference at birth | −0.06 |
| Siblings | 0.39 |
| Livestock | −0.64 |
| Family history of asthma | 0.53 |
| Family history of hay fever | 0.83 |
| Never drank tea | 0.18 |
| Nonparent childcare | −0.97 |
| Baby vaccines | 0.99 |
| Outside hours | −0.07 |

We calculated the AUC using 10-fold cross-validation to evaluate prediction accuracy. For each model, the AUC is calculated using the 5 imputed datasets separately and the results are combined and averaged.

Table 7 shows the AUC values of each data and the mean values when fitting Model 1 and 2, and presents the average sensitivity and specificity of the five datasets. Figure 3 shows the ROC curves of the two models (Model 1 and Model 2). The curves are the ROC curves for each dataset, and it can be concluded that the ROC curves obtained by five imputed datasets with the same model are very close. From the AUC results we can conclude that neither of the two prediction models is very accurate on this dataset. In order to combine the results of the analysis, the coefficient estimates from the five same models are averaged.

**Table 7:** AUC, sensitivity and specificity of models for asthma.

| Model | AUC |  |  |  |  |  | Sensitivity (%) | Specificity (%) |
| --- | --- | --- | --- | --- | --- | --- | --- | --- |
|  | 1 | 2 | 3 | 4 | 5 | Mean | Mean | Mean |
| Model 1 | 0.68 | 0.66 | 0.67 | 0.66 | 0.67 | 0.67 | 68 | 63 |
| Model 2 | 0.65 | 0.67 | 0.66 | 0.66 | 0.66 | 0.66 | 76 | 52 |
| Model 1 + LCI | 0.68 | 0.66 | 0.67 | 0.65 | 0.66 | 0.66 | 63 | 66 |
| Model 1 + FRC | 0.68 | 0.66 | 0.67 | 0.65 | 0.66 | 0.66 | 65 | 65 |
| Model 1 + CEV | 0.68 | 0.66 | 0.67 | 0.66 | 0.66 | 0.67 | 64 | 66 |
| mpsmoke | 0.38 | 0.37 | 0.39 | 0.37 | 0.4 | 0.38 | 100 | 1 |
| Model 1 + mpsmoke | 0.66 | 0.64 | 0.65 | 0.64 | 0.64 | 0.65 | 66 | 60 |
| Model 2 + mpsmoke | 0.64 | 0.63 | 0.64 | 0.62 | 0.63 | 0.63 | 54 | 71 |
| anympsmoke | 0.44 | 0.44 | 0.45 | 0.42 | 0.45 | 0.44 | 18 | 86 |
| Model 1 + anympsmoke | 0.68 | 0.66 | 0.67 | 0.65 | 0.66 | 0.66 | 64 | 65 |
| Model 2 + anympsmoke | 0.66 | 0.65 | 0.66 | 0.64 | 0.65 | 0.65 | 69 | 58 |

**Figure 3:**
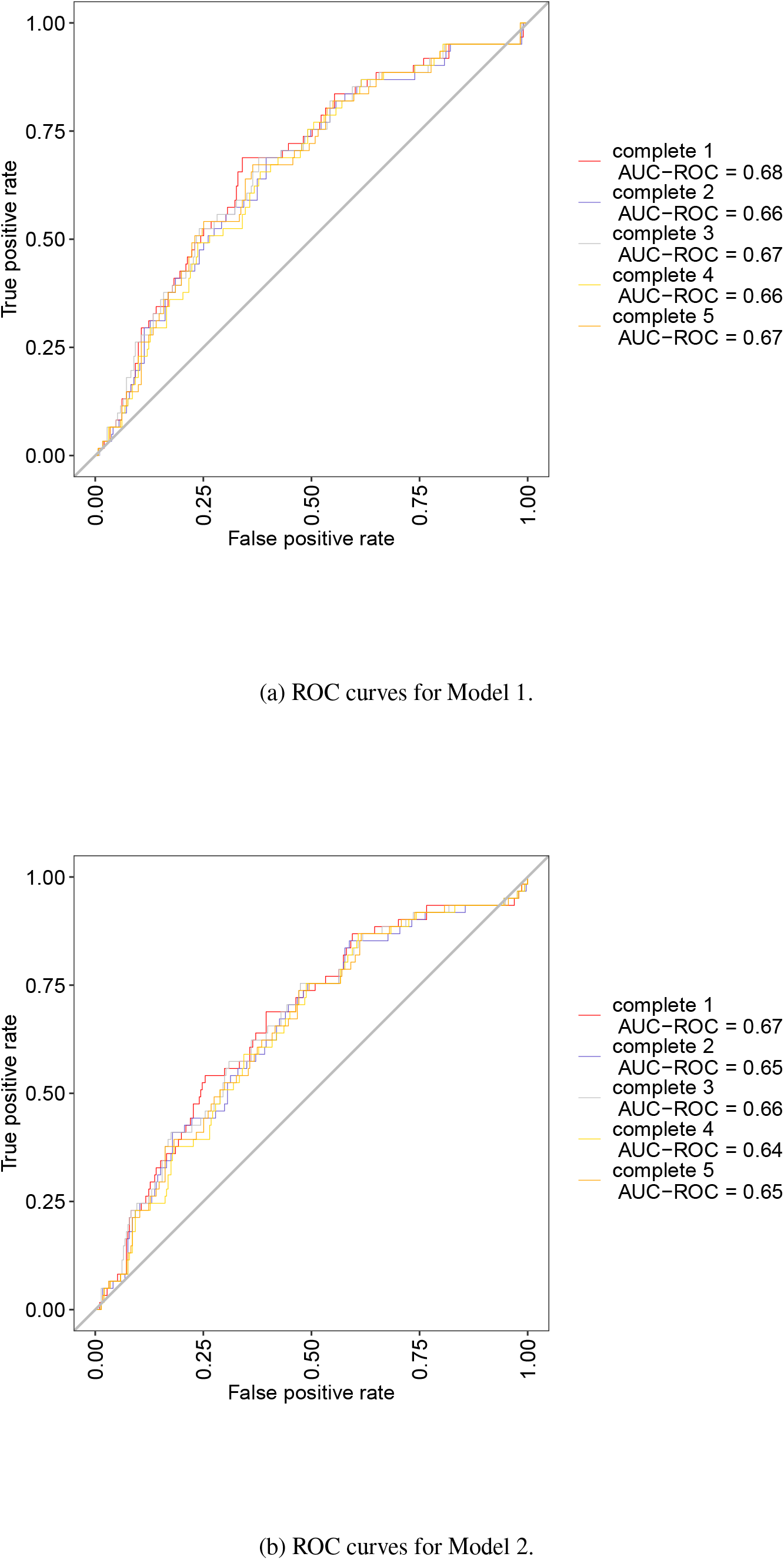
ROC curves for Model 1 and Model 2.

#### 3.1.2 Exploring the predictive power of lung function measurements

Logistic regression analysis has previously been applied to predict the risk of respiratory diseases. Notably, in many similar studies, the prediction models included maternal smoking variables as well as lung function indices. However, according to the results in the previous sections, neither stepwise selection nor LASSO methods for variable selection included `mpsmoke` or `anympsmoke`, variables that were concluded to be important in previous studies, in the models. Although the LCI variables were included in Model 2, the predictive model did not perform well. These variables do not appear to be important predictor variables in this case.

To explore the predictive power of LCI, CEV and FRC in the models, each variable was individually placed into the model with the outcome `asthma_4y` to see the accuracy of the predictive model. Table 8 provides the AUC for the three univariate models.

**Table 8:** AUC for models with only MBW measurements, for asthma.

| Model | AUC |
| --- | --- |
| LCI | 0.48 |
| FRC | 0.32 |
| CEV | 0.39 |

Using the models (Model 1) obtained in Section 3.1, the three variables of MBW were added separately to see if the accuracy of the models improved. This could help us determine if the variable could be an important factor in predicting asthma in this case. Table 7 shows a comparison of the results of AUC after adding a lung respiratory parameter variable to the original model.

From the above figures and tables, it can be concluded that the variables LCI, FRC, and CEV do not have the ability to predict the risk of having asthma at 4 years of age. When one of the MBW variables is fitted to the model alone, the AUC is below 0.5. When they are added individually to Model 1, the AUC of the new model does not increase. Sensitivity and specificity of the new model are also described in Table 7 to facilitate comparisons between models.

The results showed that the predictive ability of the model including the MBW variables also did not improve significantly, and to explore the reason, the relationship between these variables and the response variable was plotted separately. Specifically, the variables within the distribution of the data on the *x*-axis are evenly and reasonably divided into n intervals, the proportion of observations with `asthma_4y` in each interval is calculated, and each interval is plotted with this proportion and its confidence interval (CI) to determine from the distribution of the data, and the calculation whether there is a trend in this variable with respect to asthma. The confidence interval (CI) is the overall range of values that may be included for a given level of confidence. As the sample size increases, the range of interval values will become narrower, meaning that the mean has a higher degree of accuracy than a smaller sample.

Figure 4 shows the relationship between each of the three lung function parameters and the response variable `asthma_4y`. As can be seen in Figure 4a, the LCI data is concentrated between 5.5 and 6.75, with the majority of subjects not having asthma. The proportion in each interval fluctuates between 0.2 and 0.3 and does not show a very clear trend. Also, because observations in the first and last interval did not have asthma, no confidence intervals are shown. The numbers in the second interval are smaller and the confidence interval range is large.

**Figure 4:**
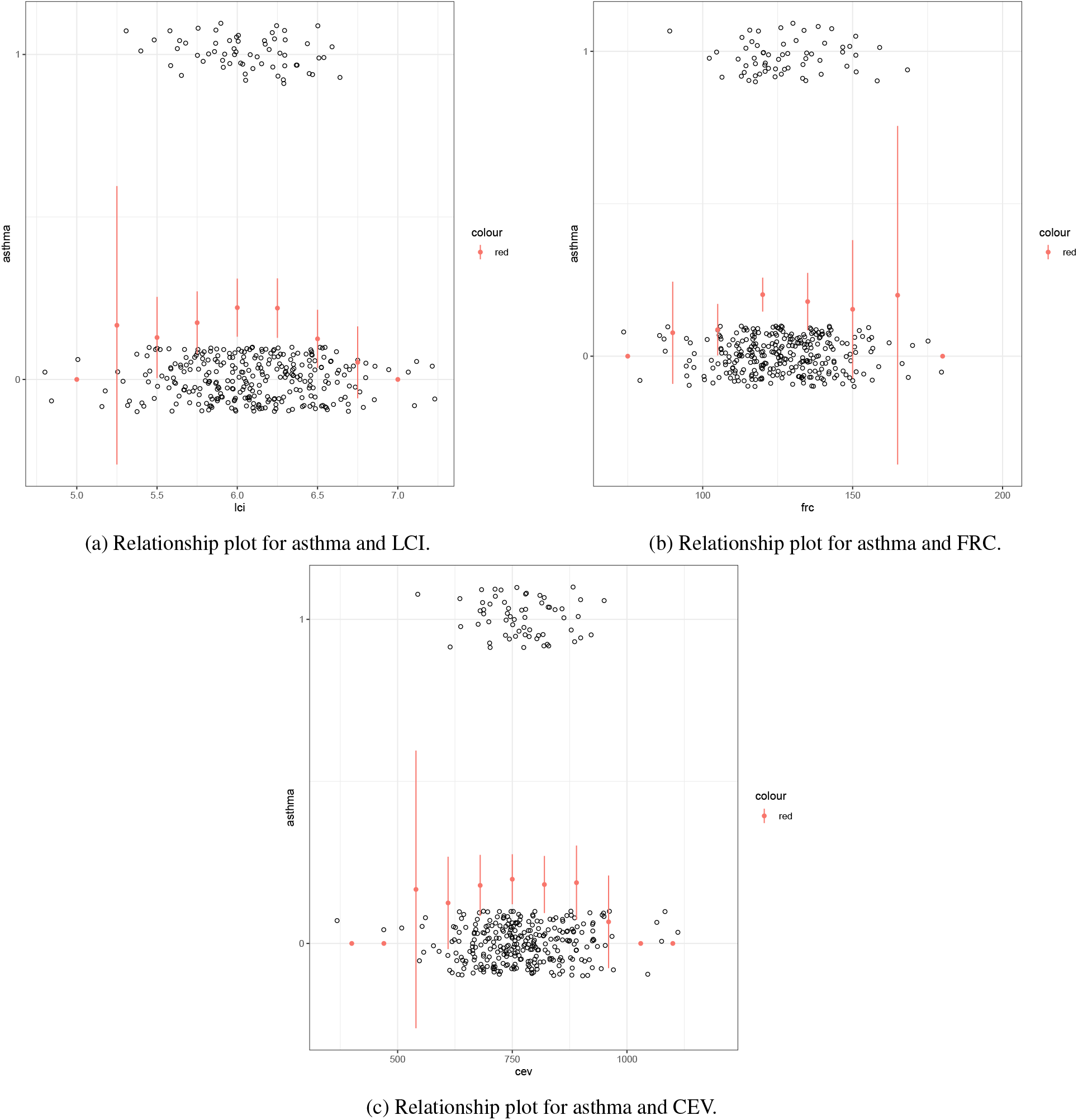
The relationship between asthma and MBW data.

Similarly, the Figure 4 also shows that the other two variables, FRC and CEV, do not have a clear trend of being related to the outcome variable, i.e. the proportions do not change significantly as the values of the two variables increase. We can observe a weak U-shaped curve between the MBW data and the asthma; we did not attempt to model this relationship.

#### 3.1.3 Exploring the predictive power of smoking

Another variable of interest is `mpsmoke`, which is a variable with six categories that represent the severity of the mother’s smoking during pregnancy. To explore the utility of this for the prediction of the 4-year-old phenotypes, we followed the same approach as in the previous section, for examining the lung function measurements.

The AUC of the relevant models are presented in the Table 7. Similar to the previous section, our results indicated that the smoking variable clearly did not help to improve prediction accuracy.

### 3.2 Wheeze

We followed the same process as for asthma and got similar results. The response variable in this case was `wheeze_4y`. Table 9 and Table 10 show the selected variables and their average fitted coefficients.

**Table 9:** Estimated coefficients for Model 3.

| Variable | Coefficient |
| --- | --- |
| Tricep skin fold at birth | 0.37 |
| Subscapular fold at birth | −0.22 |
| Family history of asthma | 0.67 |
| Family history of eczema | 0.84 |

**Table 10:** Estimated coefficients for Model 4.

| Variable | Coefficient |
| --- | --- |
| Middle upper arm circumference at birth | 0.11 |
| Tricep skin fold at birth | 0.18 |
| Mother's age | 0.03 |
| Father's age | 0.02 |
| Siblings | 0.14 |
| Livestock | −0.48 |
| Family history of asthma | 0.72 |
| Family history of eczema | 0.74 |
| Family history of hay fever | 0.26 |

The performance metrics for Model 3 and Model 4 are presented in Table 11. Model 3 has better predictive ability, but not to a very high level, with a mean AUC of 0.65.

**Table 11:**
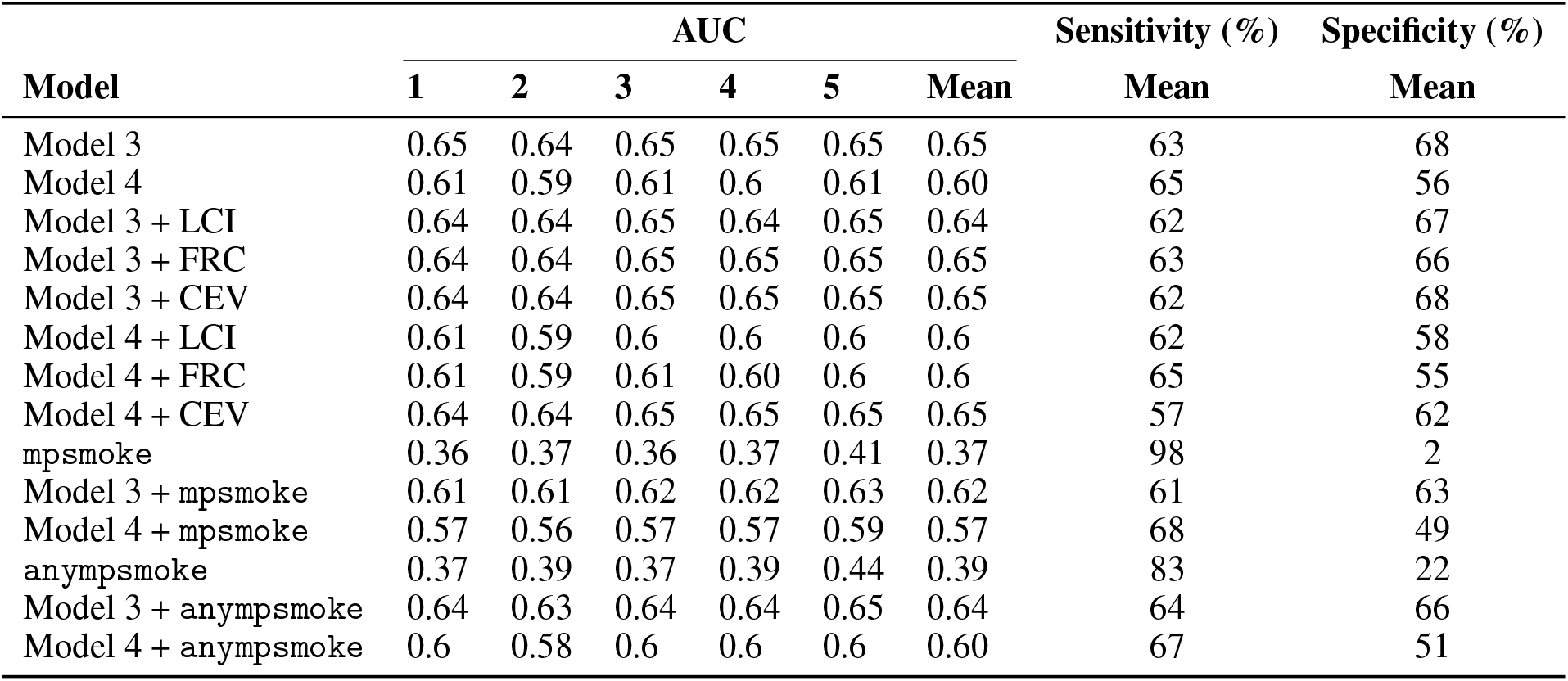
AUC, sensitivity and specificity of models for wheeze.

**Table 12:** AUC for models with only MBW measurements, for wheeze.

| Model | AUC |
| --- | --- |
| LCI | 0.39 |
| FRC | 0.42 |
| CEV | 0.44 |

We followed the same process as for asthma to explore the utility of the lung function measurements and maternal smoking during pregnancy. The results are summarized in Table 11. The conclusions are the same: these specific variables did not improve prediction accuracy.

### 3.3 Hay fever

We repeated the same analyses but with hay fever as the outcome variable. Table 13 and Table 14 show the selected variables and their average fitted coefficients. Table 15 provides the main results.

**Table 13:** Estimated coefficients for Model 5.

| Variable | Coefficient |
| --- | --- |
| Mother's age | 0.12 |
| Father's age | −0.15 |
| Family history of hay fever | 0.96 |
| Never drank tea | 1.01 |
| Pesticides | 0.95 |
| Baby handwash frequency_2 | 15.20 |
| Baby handwash frequency_3 | 13.50 |
| Baby handwash frequency_4 | 12.61 |

**Table 14:** Estimated coefficients for Model 6.

| Variable | Coefficient |
| --- | --- |
| Father's age | −0.08 |
| Family history of hay fever | 0.86 |
| Family history of eczema | 0.44 |
| Never drank tea | 0.92 |
| Pesticides | 0.87 |
| Baby handwash frequency_2 | 15.25 |
| Baby handwash frequency_3 | 13.7 |
| Baby handwash frequency_4 | 12.86 |

**Table 15:** AUC, sensitivity and specificity of models for hay fever.

| Model | AUC |  |  |  |  |  | Sensitivity (%) | Specificity (%) |
| --- | --- | --- | --- | --- | --- | --- | --- | --- |
|  | 1 | 2 | 3 | 4 | 5 | Mean | Mean | Mean |
| Model 5 | 0.71 | 0.7 | 0.71 | 0.71 | 0.68 | 0.70 | 68 | 67 |
| Model 6 | 0.69 | 0.67 | 0.7 | 0.69 | 0.66 | 0.68 | 79 | 58 |
| Model 5 + LCI | 0.7 | 0.69 | 0.71 | 0.7 | 0.68 | 0.70 | 71 | 64 |
| Model 5 + FRC | 0.7 | 0.69 | 0.71 | 0.7 | 0.68 | 0.70 | 69 | 66 |
| Model 5 + CEV | 0.7 | 0.69 | 0.71 | 0.7 | 0.68 | 0.70 | 74 | 61 |
| Model 6 + LCI | 0.69 | 0.67 | 0.69 | 0.68 | 0.66 | 0.68 | 79 | 57 |
| Model 6 + FRC | 0.69 | 0.67 | 0.69 | 0.68 | 0.66 | 0.68 | 79 | 58 |
| Model 6 + CEV | 0.7 | 0.69 | 0.67 | 0.768 | 0.68 | 0.69 | 78 | 58 |
| mpsmoke | 0.44 | 0.4 | 0.42 | 0.44 | 0.44 | 0.43 | 95 | 11 |
| Model 5 + mpsmoke | 0.7 | 0.7 | 0.71 | 0.71 | 0.68 | 0.70 | 74 | 63 |
| Model 6 + mpsmoke | 0.69 | 0.67 | 0.7 | 0.68 | 0.66 | 0.68 | 77 | 59 |
| anympsmoke | 0.42 | 0.4 | 0.4 | 0.4 | 0.42 | 0.41 | 68 | 35 |
| Model 5 + anympsmoke | 0.69 | 0.69 | 0.7 | 0.7 | 0.67 | 0.69 | 68 | 67 |
| Model 6 + anympsmoke | 0.68 | 0.66 | 0.68 | 0.68 | 0.65 | 0.67 | 74 | 58 |

**Table 16:** AUC for models with only MBW measurements, for hay fever.

| Model | AUC |
| --- | --- |
| LCI | 0.41 |
| FRC | 0.33 |
| CEV | 0.44 |

We followed the same process as for asthma to explore the utility of the lung function measurements and maternal smoking during pregnancy. The results are summarized in Table 15. The conclusions are the same: these specific variables did not improve prediction accuracy.

## 4 Discussion

We analysed the BIS data to assess our capability to predict asthma, wheeze and hay fever in children of four years of age, using lung function and other measurements obtained at birth. Our general conclusions, outlined below, applied similarly to all three diseases that we studied.

### Predictive performance is modest, and on par with existing literature

Using logistic regression, together with either stepwise selection or LASSO, our main conclusion was that our models have only a modest ability to predict the presence of the three diseases. For the best models, the estimated AUC was typically less than 0.7, and the sensitivity and specificity was typically less than 70%. We therefore expect that these models are unlikely to be useful in a clinical setting.

While these results might not yet reach a strong clinical standard, we note that this is a challenging prediction problem and that we compare favourably with other similar attempts. By way of comparison, Castro-Rodríguez et al. (2000) created a clinical index using children with wheezing symptoms in the previous three years to predict the risk of having asthma at ages 6, 8, 11 and 13. The model had a sensitivity of 56.6% and a specificity of 80.8% in predicting disease at age of 6 years. This is a less challenging scenario than ours: by the age of 3 there is substantially more medical information to work with, and they aim to predict only 3 years ahead rather than 4. Nevertheless, our predictive performance is only marginally worse than what they acheived. When our models for asthma are calibrated to a specificity of around 80%, we obtain a sensitvity of around 30%; conversely, we can achieve about 60% sensitivity when operating at a specificity of around 70%.

### Maternal smoking was a poor predictor

Interestingly, maternal smoking was not a good predictor in our models, despite showing a strong relationship with the risk of respiratory disease in previous literature (Carlsen et al., 1997; Hanrahan et al., 2012; Weitzman et al., 1990; Martinez et al., 1995; Stick et al., 1996).

One possible reason is that there were relatively few smokers in our study. We may have simply had insufficient information to estimate the impact of smoking.

A similar study to ours concluded that maternal smoking was associated with a high prevalence of asthma in children under 5 years of age (Weitzman et al., 1990). For that study, the mothers who smoked daily accounted for 25.9% of the total sample. For the BIS dataset that we used in our study, only 13.7% of mothers smoked at all.

### MBW-derived lung function measures were poor predictors

One of our aims was to use lung function measurements from birth to help improve prediction.

The evidence from previous studies on whether this might be successful has been mixed so far. For example, Pike et al. (2011) reported that children with low lung function at birth have a higher chance of developing wheezing at three years of age, however Martinez et al. (1991) also showed that wheezing at six years of age does not seem to be associated with low lung function in children. Proietti et al. (2014) also found a weak correlation between tidal breathing parameters and subsequent respiratory symptoms.

Unfortunately, in our study, the inclusion of any of the three MBW variables (LCI, FRC and CEV) did not provide any noticeable improvement in predicting the three diseases we studied at 4 years of age.

### Family history was the best predictor

Of the other variables, ones that were consistently selected for inclusion were those that related to family history of respiratory diseases. This is reassuring, given that family history is known to be a good predictor.

#### 4.1 Limitations and future work

We highlight the following limitations of our study and suggestions for future research:

1. When MI was used to impute missing values, both the predictors and the response variables were imputed. However, when fitting models to each disease, not all 401 participants were used, but instead observations with no missing values for the response variable before imputation were selected. It is worth noting that it is reasonable to fill in the missing values for the response variable and use them in the data analysis (Steyerberg et al., 2019). The data after including these participants could be reanalyzed in a subsequent study, to see whether and how the results change.
2. We used two methods (stepwise selection and LASSO) to select the variables and then used the selected model to obtain the parameters of the model using the cross-validation method, but the results obtained following this step will be overoptimistic. Doing cross-validation after model selection will result in an overestimate of the predictive performance. A better way would be to do the CV with the whole model fitting process, i.e. perform the variable selection within each fold. The extent of any overestimation should be assessed in a future project.
3. We can observe weak U-shaped curves in the relationship between certain MBW variables and the presence of the respiratory diseases. This suggests exploring non-linear relationships in the modellig. For example, adding quadratic terms, such as LCI^2^, to the previously selected models to see if the predictive power of the model is improved.

## Supporting information

Tripod checklist

## Data Availability

For queries about the data, please contact the Barwon Infant Study at

